# Towards harmonizing subtyping methods for neuroimaging studies in Alzheimer’s disease

**DOI:** 10.1101/2020.04.19.20064881

**Authors:** Rosaleena Mohanty, Gustav Mårtensson, Konstantinos Poulakis, J-Sebastian Muehlboeck, Elena Rodriguez-Vieitez, Konstantinos Chiotis, Michel J. Grothe, Agneta Nordberg, Daniel Ferreira, Eric Westman

## Abstract

**Background:** Biological subtypes in Alzheimer’s disease (AD), originally identified on neuropathological data, have been translated to *in vivo* biomarkers such as structural magnetic resonance imaging (sMRI) and positron emission tomography (PET), to disentangle the heterogeneity within AD. Although there is methodological variability across studies, comparable characteristics of subtypes are reported at the group level. In this study, we investigated whether group-level similarities translate to individual-level agreement across subtyping methods, in a head-to-head context.

**Methods:** We compared five previously published subtyping methods. Firstly, we validated the subtyping methods in 89 amyloid-beta positive (Aβ+) AD dementia patients (reference group: 70 Aβ-healthy individuals; HC) using sMRI. Secondly, we extended and applied the subtyping methods to 53 Aβ+ prodromal AD and 30 Aβ+ AD dementia patients (reference group: 200 Aβ-HC) using both sMRI and tau PET. Subtyping methods were implemented as outlined in each original study. Group-level and individual-level comparisons across methods were performed.

**Results:** Each individual method was replicated and the proof-of-concept was established. All methods captured subtypes with similar patterns of demographic and clinical characteristics, and with similar maps of cortical thinning and tau PET uptake, at the group level. However, large disagreements were found at the individual level.

**Conclusions:** Although characteristics of subtypes may be comparable at the group level, there is a large disagreement at the individual level across subtyping methods. Therefore, there is an urgent need for consensus and harmonization across subtyping methods. We call for establishment of an open benchmarking framework to overcome this problem.

## 1. INTRODUCTION

The study of biological subtypes has opened a great opportunity to unravel the heterogeneity within Alzheimer’s disease (AD). The topic was rekindled in 2011 by the seminal study from Murray et al. (1), and during the last five years it has exploded with numerous structural magnetic resonance imaging (sMRI) subtyping studies (see (2) for a review). In 2018, the first tau positron emission tomography (PET) subtyping study was published (3), and more are expected to come in the near future.

However, studies investigating AD subtypes differ considerably, with almost no methodological consensus. Murray et al. (1) based subtyping on postmortem tau neurofibrillary tangle (NFT) counts in the hippocampus and three cortical regions. All patients were at Braak’s stage V or VI (4) and were classified into three subtypes according to the 25^th^ and 75^th^ percentiles in the hippocampus-to-cortex index: *typical AD, limbic-predominant AD*, and *hippocampal-sparing AD*. Byun et al. (5) translated this subtyping method to sMRI data with volumes of the same brain regions as in Murray’s method, but defined abnormality as −1 standard deviation from age-, sex-, and intracranial volume (ICV)-adjusted normative data of healthy controls. This method identified a fourth subtype: *minimal atrophy AD* (5). In contrast, Risacher et al. (6) followed the 25^th^ and 75^th^ percentiles procedure using the hippocampus-to-cortex index, but with seven cortical regions extending the three regions used in Murray et al. (1). Risacher et al. (6) also corrected for age, sex, and ICV, but they based this correction on the amyloid-beta negative (Aβ-) healthy controls as the reference group, and used a different correction method that also included the MRI field strength. Studies from our lab used visual rating scales of brain atrophy in medial temporal, frontal, and posterior cortices (7–13), and determined abnormality based on clinical cut points (14). We also used an unsupervised clustering method in another cross-sectional study (15), which has recently been extended for subtyping on longitudinal data (16). Other groups used different unsupervised clustering methods (17–24), highlighting the methodological variability across studies. Additionally, Charil et al. (25) recently translated Murray’s method to tau PET while Whitwell et al. (3) applied a clustering method on tau PET data.

Despite this variability, all these studies tend to identify subtypes with similar characteristics, arguing for validation (see (2) for a review). However, this validation is reported at the group level. The ultimate goal of investigating heterogeneity in AD is to understand individual variability; hence validation also needs to be demonstrated at the individual level. Surprisingly, no head-to-head comparison of subtyping methods has been published so far. Such a comparison arises as an urgent and important step towards facilitating consistent progress in this field, especially with the current surge in subtyping studies using longitudinal sMRI (16,26,27) and tau PET (3,25,28). To illustrate this problem and substantiate our claim for harmonizing subtyping methods, we applied subtyping methods based on five previous studies (1,5–7,15,25), on sMRI and tau PET data, in the same cohort. In our primary analyses, we performed a head-to-head comparison and report subtypes’ frequencies, characteristics, and cortical thickness and tau PET uptake maps from the different methods. In our secondary analyses, we investigated how methodological variations influence the performance of the different subtyping methods. We hypothesized that the different methods would provide largely comparable subtypes at the group level, while disagreement would be larger at the individual level.

## 2. METHODS

### 2.1. Participants

All participants were selected from the ADNI (http://adni.loni.usc.edu/). The goal of the ADNI (launched in 2003, PI: Michael W. Weiner; (29)) is to measure the progression of prodromal AD and early AD using MRI, PET, and biomarkers, as well as clinical and neuropsychological assessments. Two separate ADNI cohorts were included in this study:

Firstly, given that subtypes were originally identified in AD dementia, we validated the previously published subtyping methods using sMRI in a cohort of 89 AD dementia patients (Aβ+) from ADNI-1. We also included a control group of 70 Aβ-healthy individuals (HC). Amyloid status was determined on the basis of cerebrospinal fluid biomarkers (30).

Secondly, subtyping was applied to a cross-sectional cohort of 84 patients (54 Aβ+ prodromal AD patients, 30 Aβ+ AD dementia patients) from ADNI-2 and −3 using both sMRI and tau PET. The control group for this cohort comprised 200 Aβ-HC. Here, amyloid status was determined through amyloid PET (florbetapir cut-off = 1.11; (31) or florbetaben cut-off = 1.08 from ADNI’s current recommendation, http://adni.loni.usc.edu/).

We will refer to these two cohorts as the *sMRI cohort* (ADNI-1, AD dementia patients) and the *sMRI-tauPET cohort* (ADNI-2 and −3, prodromal AD and AD dementia patients). We validated the previously published methods in the sMRI cohort and extended our analyses to the sMRI-tauPET cohort. The study protocol followed by all participating centers within the ADNI was approved by their respective institutional review board. Informed and written consent was obtained from all the participants.

### 2.2. MRI and PET imaging

#### MRI acquisition and processing

3-D accelerated T1-weighted sequences were acquired with sagittal slices and voxel size 1.1 × 1.1 × 1.2 mm^3^. MRI data for the ADNI-1 were acquired on 1.5T scanners, and MRI data for ADNI-2 and −3 were acquired on 3.0T scanners.

For the sMRI cohort, processed data were already available from our previous studies (7,15). For methods from other labs (5,6) and for all the methods in the sMRI-tauPET cohort, data were unavailable, so we processed the sMRI through TheHiveDB system (32) with FreeSurfer 6.0.0 (http://freesurfer.net/).Following the cross-sectional stream, quality control of the output from FreeSurfer was conducted visually. Automatic region of interest parcellation yielded volumetric measures for cortical and subcortical brain structures (33–35). For the subtyping method using visual rating scales (7), the rating scales were computed automatically using AVRA (Automatic Visual Ratings of Atrophy) v0.8 (https://github.com/gsmartensson/avra_public) (36). AVRA is a deep learning model trained on over 3000 MRI scans rated by an expert neuroradiologist with excellent inter-rater agreement (37).

#### Tau PET acquisition and processing

Tau PET scans were collected using a GE PET/CT scanner. [^18^F]AV-1451 was injected with a dosage of 370 MBq (10.0 mCi) ± 10% and scans were acquired between 75-105 min post-injection. The dynamic acquisition was 30 min long and comprised of 6 × 5 min frames. For each tau PET scan, a sMRI scan was available within 90 days (except in 3 AD dementia and 5 prodromal AD patients, >90 days).

For subtyping methods using tau PET (1,5,6,25), processing was performed using the PetSurfer Toolbox (38) within FreeSurfer 6.0.0. AV-1451 images were co-registered onto the corresponding FreeSurfer-processed sMRI. The regions (cortical and subcortical) estimated for each individual were consistent with those used for sMRI-based subtypes (33). Partial volume correction (PVC) was applied using the symmetric geometric matrix method (39). AV-1451 signal was quantified in each region as the standardized uptake value ratio (SUVR), computed with the cerebellum grey matter as the reference region with PVC.

### 2.3. Subtyping methods

Based on two recent systematic reviews (2,40), we identified four sources of methodological variation in previous subtyping studies:

i. *Type of method* (hypothesis-driven *vs*. data-driven).
ii. *Definition of subtype* (dependent on the sample of study *vs*. dependent on an external reference group).
iii. *Modality* (postmortem NFT *vs*. sMRI *vs*. tau PET).
iv. *Measure* (NFT count *vs*. automated volumes *vs*. automated SUVR values *vs*. visual ratings).

The method proposed by Murray et al. (1) is the only one based on postmortem NFT count and motivated subsequent neuroimaging studies. In this study, we focused on neuroimaging-based methods based on five subtyping studies, covering all these levels of methodological variation: Risacher et al. (6), Byun et al. (6), Ferreira et al. (7), Poulakis et al. (15), and Charil et al. (25). Each subtyping method was implemented to replicate the original method as closely as possible, as elaborated further in **Table 1 and Supplemental Table S1**. We also translated some of the sMRI-based methods to tau PET (5,6). For Byun’s method on tau PET, we identified a minimal tau subtype that is not captured by Charil’s or Risacher’s methods.

**Table 1.**
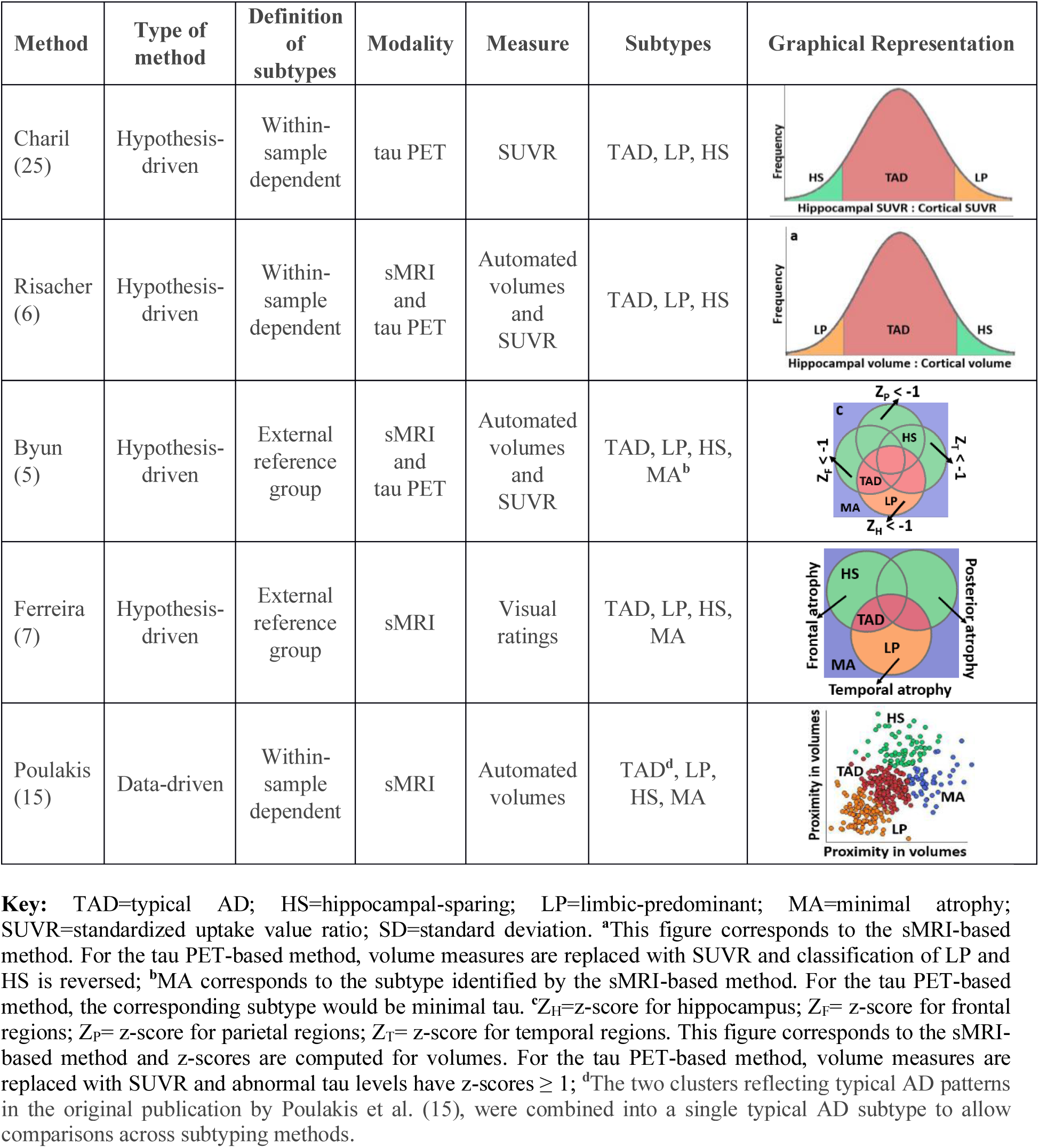
Overview of the subtyping methods implemented in this study.

| Method | Type of method | Definition of subtypes | Modality | Measure | Subtypes | Graphical Representation |
| --- | --- | --- | --- | --- | --- | --- |
| Charil (25) | Hypothesis-driven | Within-sample dependent | tau PET | SUVR | TAD, LP, HS |  |
| Risacher (6) | Hypothesis-driven | Within-sample dependent | sMRI and tau PET | Automated volumes and SUVR | TAD, LP, HS |  |
| Byun (5) | Hypothesis-driven | External reference group | sMRI and tau PET | Automated volumes and SUVR | TAD, LP, HS, MA<sup>b</sup> |  |
| Ferreira (7) | Hypothesis-driven | External reference group | sMRI | Visual ratings | TAD, LP, HS, MA |  |
| Poulakis (15) | Data-driven | Within-sample dependent | sMRI | Automated volumes | TAD<sup>d</sup>, LP, HS, MA |  |

**Key:** TAD=typical AD; HS=hippocampal-sparing; LP=limbic-predominant; MA=minimal atrophy; SUVR=standardized uptake value ratio; SD=standard deviation. <sup>a</sup>This figure corresponds to the sMRI-based method. For the tau PET-based method, volume measures are replaced with SUVR and classification of LP and HS is reversed; <sup>b</sup>MA corresponds to the subtype identified by the sMRI-based method. For the tau PET-based method, the corresponding subtype would be minimal tau. <sup>c</sup> $Z_H$ =z-score for hippocampus; $Z_F$ = z-score for frontal regions; $Z_P$ = z-score for parietal regions; $Z_T$ = z-score for temporal regions. This figure corresponds to the sMRI-based method and z-scores are computed for volumes. For the tau PET-based method, volume measures are replaced with SUVR and abnormal tau levels have z-scores $\geq 1$ ; <sup>d</sup>The two clusters reflecting typical AD patterns in the original publication by Poulakis et al. (15), were combined into a single typical AD subtype to allow comparisons across subtyping methods.

Quantification of AV-1451 signal in the hippocampus, a key region for subtyping in many studies (5,6,25), is contentious (41,42). Hence, we additionally applied subtyping using the entorhinal cortex instead of the hippocampus, also facilitating comparability with the study by Whitwell et al. (3).

### 2.4. Methodological variations

As a secondary objective, we implemented the following methodological variations to evaluate their potential impact on agreements among subtyping methods:

#### i. The effect of using three vs. seven cortical regions in Risacher’s method

Although Risacher et al. (6) translated Murray’s method (1) to sMRI, Risacher’s method included seven cortical regions instead of the original three regions in Murray’s method. Here, we compared these two versions of Risacher’s method: with three *vs*. seven cortical regions.

#### ii. The effect of statistical corrections for ICV and age on sMRI methods

In our primary analysis, we evaluated the method by Risacher et al. (6) (seven cortical regions) by adjusting for ICV and age using a single regression model for both covariates. Here, we evaluated the impact of adjusting for ICV only, or adjusting for ICV and age using separate regression models for each covariate. We also performed these comparisons for Risacher’s method using three cortical regions.

#### iii. The effect of statistical corrections for age on tau PET methods

In the primary analysis of tau PET-based subtyping (5,6,25), potential covariates were not accounted for. Correction for ICV is not necessary unlike in sMRI methods, but age may potentially affect tau PET SUVR (43). Here, we compared subtyping with age-corrected SUVR and uncorrected SUVR.

#### iv. The effect of PVC on tau PET-based subtyping methods

In the primary analysis, we performed PVC of tau PET SUVR for a reliable quantification, as SUVR can be affected by off-target binding, especially in the hippocampus (44,45). Here, we compared subtyping between PVC SUVR and non-PVC SUVR.

### 2.5. Statistical analysis

We compared subtyping methods at the group-level in terms of age, sex, MMSE, education, and *APOE* ε4 status. Within each subtyping method, hypothesis testing was performed to compare the distribution of subtypes with the Kruskal-Wallis test. A *p*-value ≤0.05 was deemed significant. Group-level cortical thickness and tau PET uptake maps were generated by comparing each subtype with the healthy controls. In each hemisphere, data were smoothed onto the surface using a 10 mm Gaussian kernel with a full width at half maximum. A general linear model was fitted at each vertex. All maps were visualized at *p*≤0.01 (uncorrected). Individual-level agreement among subtyping methods was quantified by Cohen’s kappa (*κ* < 0, no agreement; *κ* = 0–0.20, slight agreement; *κ* = 0.21–0.40, fair agreement; *κ* < 0.41–0.60, moderate agreement; *κ* = 0.61–0.80, substantial agreement; *κ* = 0.81–1.0, almost perfect agreement) (46).

## 3. RESULTS

**Table 2** shows the demographic and clinical characteristics for the sMRI cohort (**Table 2 a**) and the sMRI-tauPET cohort (**Table 2 b**).

**Table 2.** Demographic and clinical characteristics of the cohorts.

| (a) Validation of subtyping methods in AD dementia patients (sMRI cohort) |  |  |  |  |
| --- | --- | --- | --- | --- |
|  | HC (Aβ-) |  | AD dementia (Aβ+) | p-value |
| N | 70 |  | 89 | 0.033 |
| Sex (F,%) | 51 |  | 39 | 0.139 |
| Age (years) | 75.15 ± 5.22 |  | 74.73 ± 7.72 | 0.757 |
| Education (years) | 15.66 ± 2.65 |  | 15.16 ± 3.24 | 0.402 |
| APOE ε4 carriers (%) | 10 |  | 74 | <.0001 |
| MMSE | 29.04 ± 1.10 |  | 23.48 ± 1.87 | <.0001 |
| (b) Subtyping methods in Prodromal AD and AD dementia patients (sMRI-tauPET cohort) |  |  |  |  |
|  | HC (Aβ-) | Prodromal AD (Aβ+) | AD dementia (Aβ+) | p-value |
| N | 200 | 54 | 30 | <.0001 |
| Sex (F,%) | 59 | 48 | 50 | 0.285 |
| Age (years) | 70.45 ± 5.65 | 74.09 ± 7.34 | 77.46 ± 8.27 | <.0001 |
| Education (years) | 16.90 ± 2.31 | 15.76 ± 2.66 | 15.77 ± 2.57 | 0.002 |
| APOE ε4 carriers (%) | 22 | 61 | 53 | <.0001 |
| MMSE | 29.24 ± 1.05 | 27.48 ± 2.30 | 22.13 ± 4.23 | <.0001 |

Data are reported as mean ± standard deviation;
**Key:** HC=healthy control; AD=Alzheimer's disease; sMRI=structural MRI; PET=positron emission tomography; Aβ =amyloid-beta; F=female; MMSE=Mini-Mental State Examination; *APOE*=apolipoprotein.

### 3.1. Validation of subtyping methods in the sMRI cohort

The frequencies of the subtypes in the sMRI cohort were very similar to the frequencies reported in the original studies (5–7,15,25), suggesting we could replicate the subtyping methods (**Table 3**).

**Table 3.** Frequencies of the subtypes compared with previously published studies in the sMRI cohort.

| Subtype | Risacher |  | Byun |  | Ferreira |  | Poulakis |  |
| --- | --- | --- | --- | --- | --- | --- | --- | --- |
|  | Pub. | This Study | Pub. | This Study | Pub. | This Study | Pub.<sup>a</sup> | This Study |
| Typical AD | 69 | 69 | 59 | 55 | 51 | 52 | 69 | 66 |
| Hippocampal-sparing AD | 17 | 19 | 12 | 17 | 17 | 19 | 7 | 10 |
| Limbic-predominant AD | 14 | 12 | 19 | 21 | 17 | 18 | 4 | 1 |
| Minimal atrophy AD | - | - | 10 | 7 | 15 | 11 | 19 | 23 |

Data are reported as % and rounded to the nearest integer for readability
**Key:** AD=Alzheimer's Disease; Pub.=published study. <sup>a</sup>Frequencies of subtypes based on the ADNI cohort only, since the original study by Poulakis et al. (15), also includes another cohort.

### 3.2. Group-level comparison of subtyping methods in the sMRI and sMRI-tauPET cohorts

**Figure 1** shows that, at the group-level, the subtyping methods captured similar demographic and clinical characteristics of the subtypes in both cohorts. Typical AD was always the most frequent subtype and showed greater frequency of males and lower MMSE scores relative to the other subtypes. Limbic-predominant AD showed lower MMSE scores relative to hippocampal-sparing AD. Hippocampal-sparing AD was the subtype with lowest frequency of *APOE* ε4 carriers. Minimal atrophy/minimal tau AD included younger individuals and showed higher MMSE scores.

**Figure 1.**
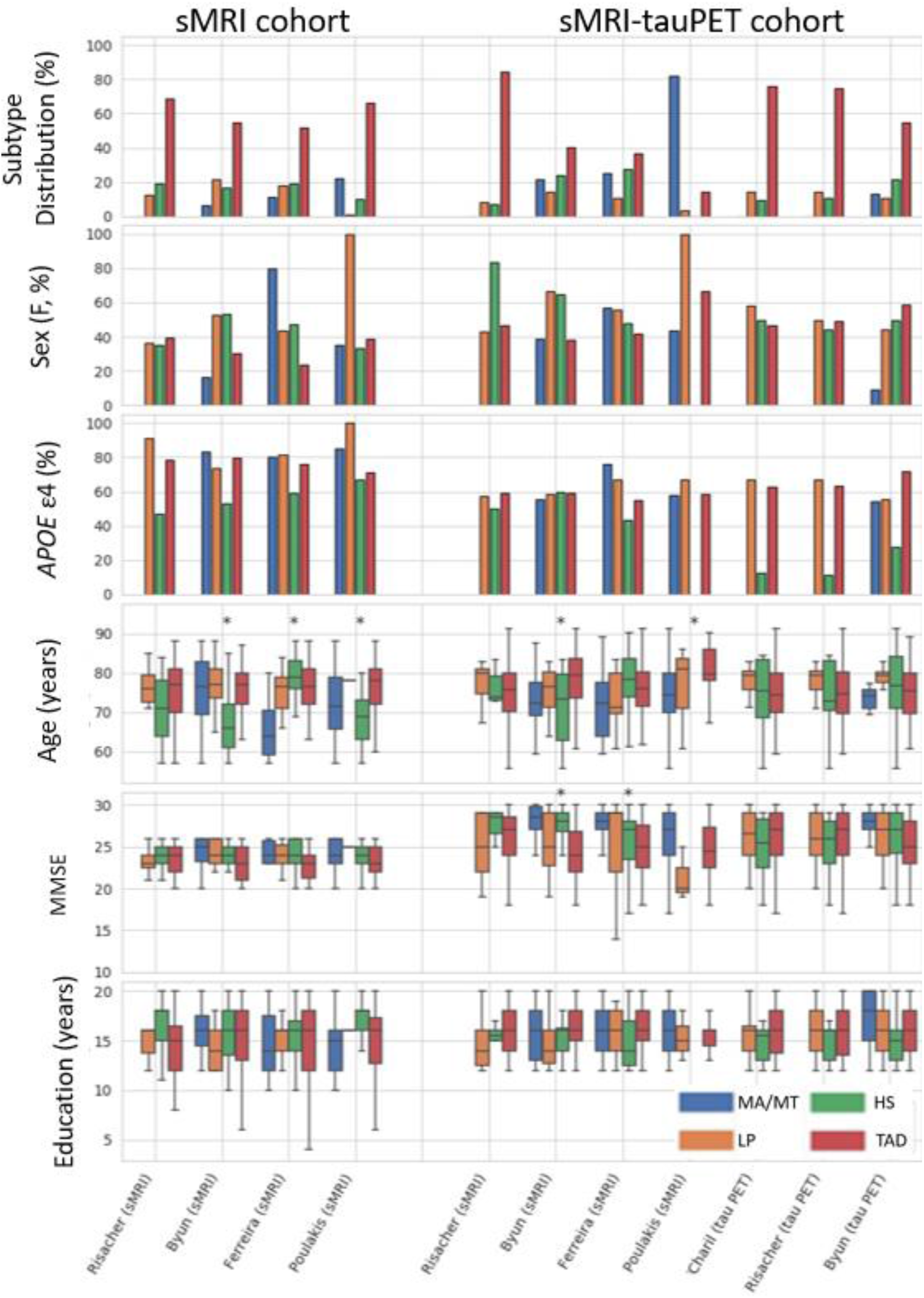
Demographic and clinical characteristics captured by the different subtyping methods. **Key:** sMRI=structural magnetic resonance imaging; PET=positron emission tomography; F=female; APOE= apolipoprotein; MMSE=mini mental state exam; MA=minimal atrophy AD; MT=minimal tau AD; LP=limbic-predominant AD; HS=hippocampal-sparing AD; TAD=typical AD.

**Figures 2-3** show that, at the group-level, the subtyping methods captured similar cortical thickness and tau PET uptake maps of the subtypes relative to healthy individuals. Cortical thinning and elevated tau PET uptake included widespread regions in typical AD; temporal and limbic regions in limbic-predominant AD; frontal or parietal regions in hippocampal-sparing AD; and relatively fewer regions in minimal atrophy/minimal tau AD, across all subtyping methods. Typical and limbic-predominant AD showed smaller hippocampal volume and greater hippocampal tau PET SUVR relative to hippocampal-sparing and minimal atrophy/minimal tau AD (boxplots in **Figures 2-3**). **Figure 4** shows the group-level tau PET uptake maps for entorhinal-based subtyping instead of hippocampus-based subtyping. Compared to hippocampus-based subtyping, albeit similar maps, hippocampal-sparing AD in entorhinal-based subtyping showed no tau PET uptake in medial temporal lobe regions. Greater tau SUVR in the entorhinal cortex was seen in typical and limbic-predominant AD than hippocampal-sparing and minimal tau AD (boxplots in **Figure 4**).

**Figure 2.**
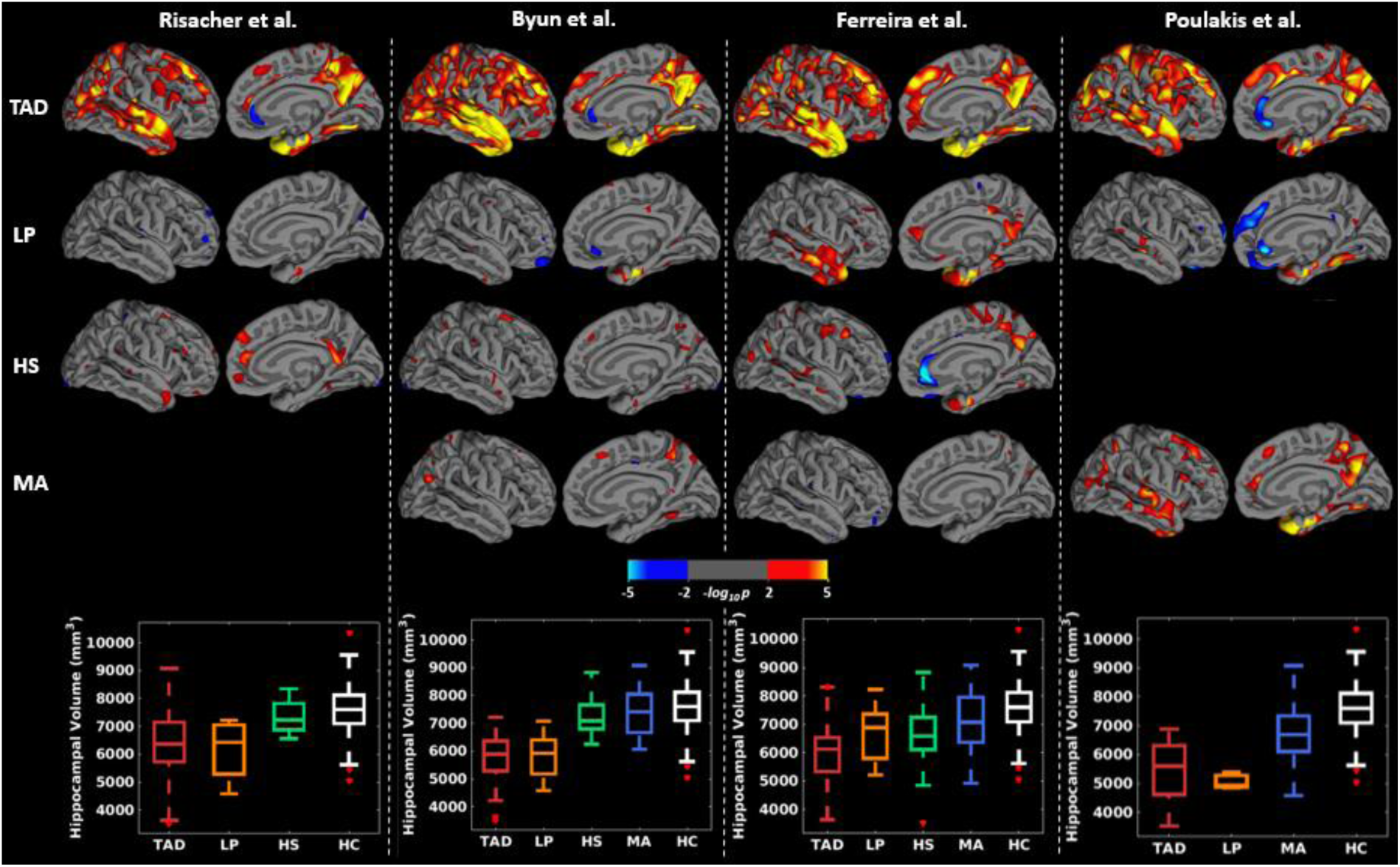
Group-level cortical thickness maps across subtyping methods in the sMRI-tauPET cohort. For simplicity, only left lateral and medial views are presented since very similar results were obtained for the right lateral and medial views. Differences in cortical thickness maps are shown in each subtype relative to HC. Yellow-red regions reflect thinner cortex in AD subtypes relative to HC. All brain maps are uncorrected for multiple comparisons at *p* < 0.01. Risacher et al. identified three subtypes only and hence, there are no cortical maps corresponding to MA subtype. Poulakis et al., identified all four subtypes. However, the HS subtype (1 individual) had to be excluded from the study due to invalid tau PET data. **Key:** TAD=typical AD; HS=hippocampal-sparing AD; LP=limbic-predominant AD; MA=minimal atrophy AD; HC=healthy control.

**Figure 3.**
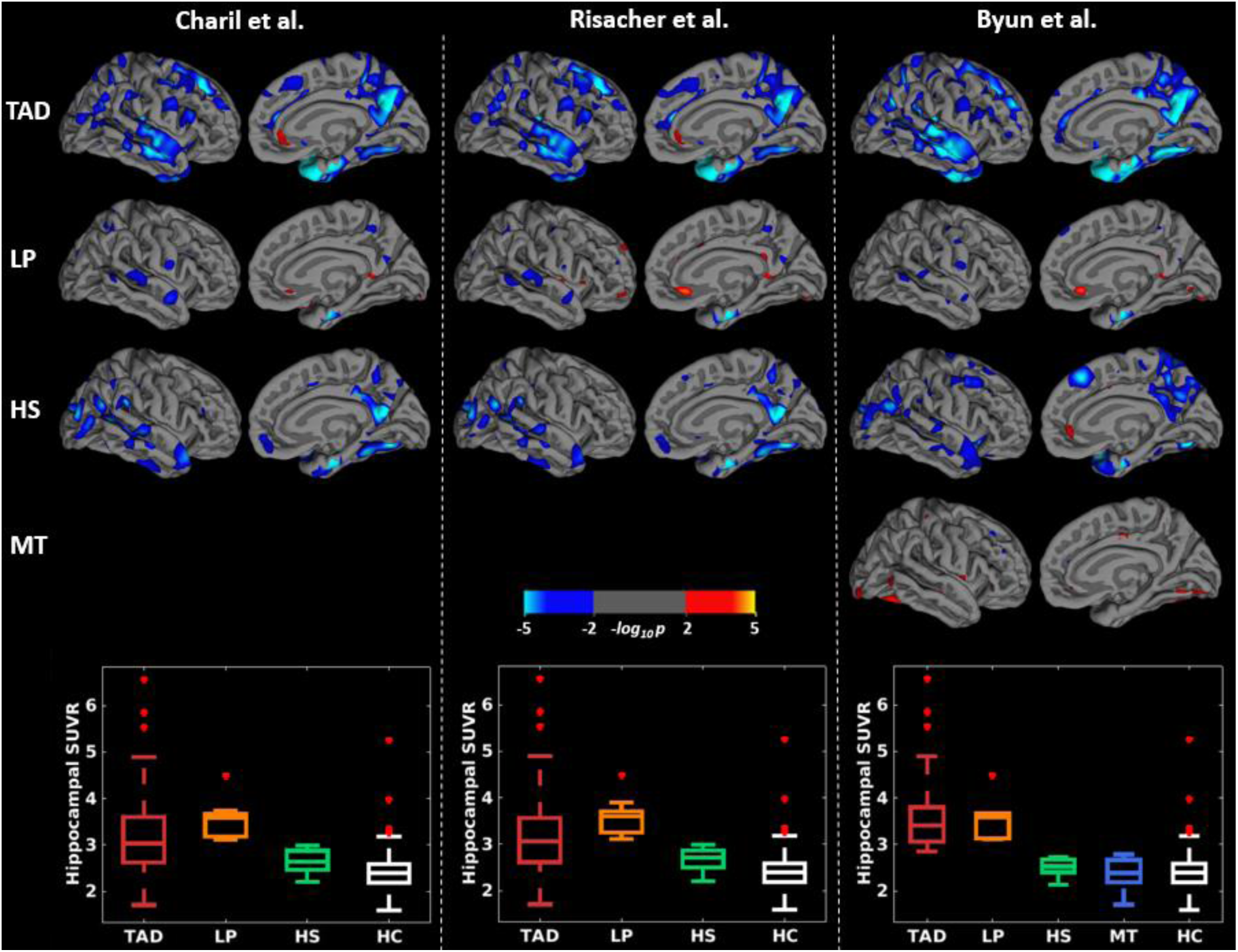
Group-level tau PET uptake maps across subtyping methods using the hippocampus in the sMRI-tauPET cohort. For simplicity, only left lateral and medial views are presented since very similar results were obtained for the right lateral and medial views. Differences in tau PET uptake maps are shown in each subtype relative to HC. Cyan regions reflect greater tau PET uptake in AD subtypes relative to HC. All brain maps are uncorrected for multiple comparisons at *p* < 0.01. **Key:** TAD=typical AD; HS=hippocampal-sparing AD; LP=limbic-predominant AD; MT=minimal tau AD; HC=healthy control.

**Figure 4.**
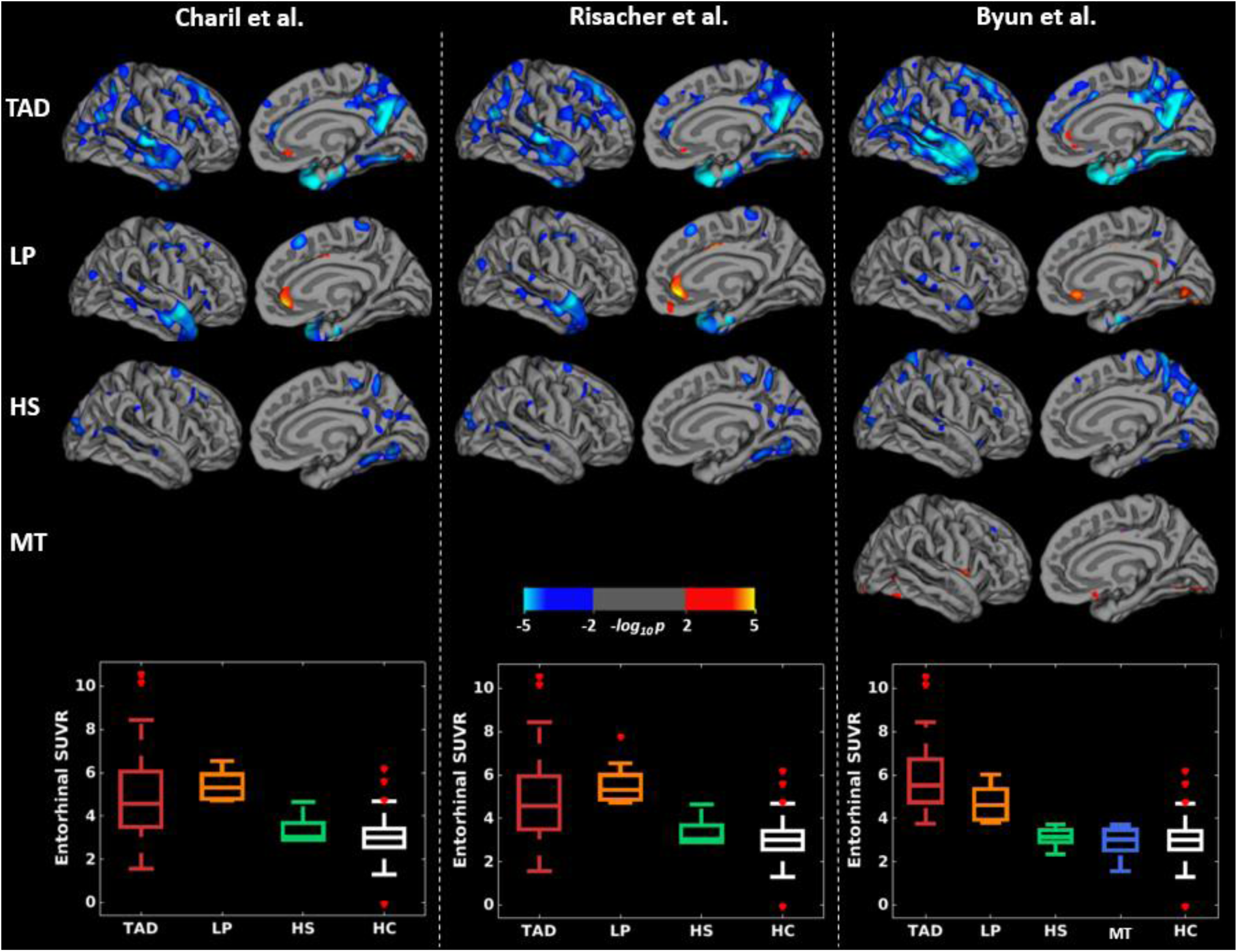
Group-level tau PET uptake maps across subtyping methods using the entorhinal cortex in the sMRI-tauPET cohort. For simplicity, only left lateral and medial views are presented since very similar results were obtained for the right lateral and medial views. Differences in tau PET uptake maps are shown in each subtype relative to HC. Blue-cyan regions reflect greater tau PET uptake in AD subtypes relative to HC. All brain maps are uncorrected for multiple comparisons at *p* < 0.01. **Key:** TAD=typical AD; HS=hippocampal-sparing; LP=limbic-predominant; MT=minimal tau; HC=healthy control.

### 3.3. Head-to-head comparison of subtyping methods in the sMRI-tauPET cohort

**Figure 5 (a, c)** shows the head-to-head comparison. Agreement among methods was low, reflected by low values of *κ*. Agreement among the tau PET-based methods was relatively higher than that of the sMRI-based methods. Since not all methods identify the minimal atrophy/minimal tau AD subtype, we excluded this subtype in follow-up analyses and observed that *κ* values increase in both cohorts and modalities (**Figure 5 b, d)**. ADNI’s participant identifiers (RID) are shown in **Figure S1** and in **Supplemental Data File**.

**Figure 5.**
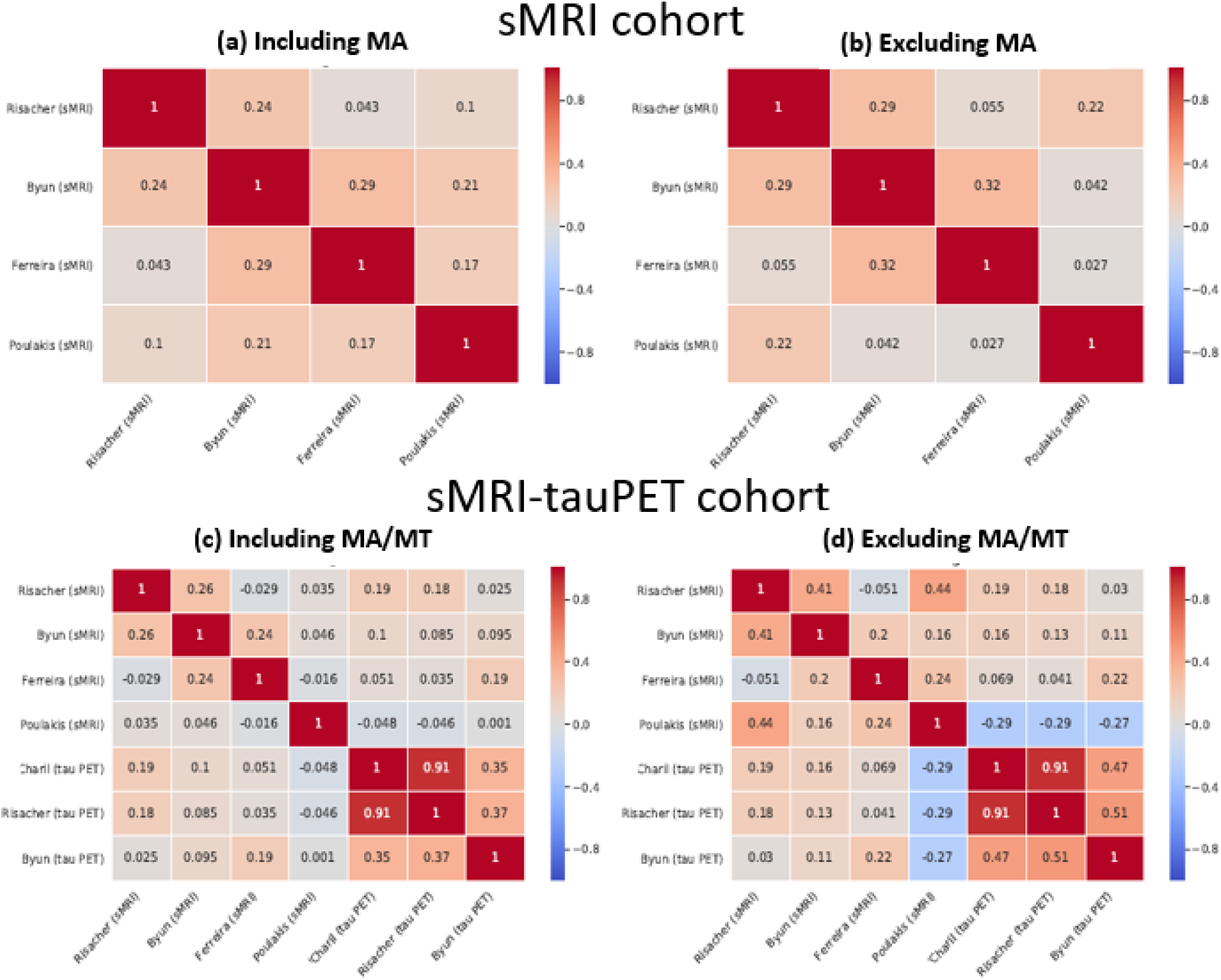
Individual-level agreement among subtyping methods as illustrated by Cohen’s kappa values. **Key:** sMRI=structural magnetic resonance imaging; PET=positron emission tomography; MA=minimal atrophy AD; MT=minimal tau.

### 3.4. Methodological variations in the sMRI-tauPET cohort

When supplementing our head-to-head comparisons with several methodological variations, we observed the following (see **Supplemental Data File**):

#### i. The effect of using three vs. seven cortical regions in Risacher’s method

Results from Risacher’s method using three cortical regions were consistent with Risacher’s method using seven cortical regions (85% agreement).

#### ii. The effect of statistical corrections for ICV and age on sMRI methods

Relative to Risacher’s method (seven cortical regions and adjusted for ICV and age in a single model), 82% of the individuals were classified consistently when performing the ICV correction only, and 69% when performing the ICV and age correction with separate models. Relative to the variation in Risacher’s method using three cortical regions (and adjusted for ICV and age in a single model), 98% of the individuals were classified consistently when performing the ICV correction only, and 74% when performing the ICV and age correction with separate models. Overall, agreements were better in typical AD (79-88%) compared to the other subtypes (15-83%).

#### iii. The effect of statistical corrections for age on tau PET methods

Over 80% of the individuals were consistently classified with and without age-adjusted tau SUVR (agreement for: Charil’s method=89%; Risacher’s method=100%; Byun’s method=80%).

#### iv. The effect of PVC on tau PET-based subtyping methods

Over 80% of the individuals were consistently classified with PVC and non-PVC SUVR (agreement for: Charil’s method=87%; Risacher’s method=89%; Byun’s method=80%). Overall, agreements were better in typical AD (83-94%) compared to the other subtypes (56-78%).

## 4. DISCUSSION

The field of biological subtypes of AD has expanded rapidly in the last decade, with numerous recent publications on neuropathological, MRI, and PET data. However, the great methodological variability is complicating reaching a definitive understanding of the heterogeneity within AD. The current study is the first head-to-head comparison of several subtyping methods in the same cohort. We found that different methods identify subtypes largely comparable at the group level (similar frequencies, demographic, clinical characteristics, cortical thinning and tau PET uptake). However, strikingly, the agreement among subtyping methods is very low when compared head-to-head at the individual level. This result may have important implications for advancing the implementation of precision medicine. Below we discuss several factors that may explain this finding and that could be addressed to minimize this problem in future studies.

Comparability across studies at the group level suggests a convergence of results and initial consensus on the existence of three-four subtypes: typical, limbic-predominant, and hippocampal-sparing subtypes in all the studies, and minimal atrophy AD in several studies. Minimal atrophy AD is only identified when considering disease severity, while the other subtypes are identified when considering typicality (2). The dimensions of severity and typicality have been identified in a recent conceptual framework for biological subtypes of AD (2). Typicality spans from limbic-predominant to hippocampal-sparing, with typical AD in-between. Severity differentiates minimal atrophy from typical AD, accounting for neurodegeneration.

The seminal study by Murray et al. (1) based subtyping on tau NFT in the hippocampus and three cortical regions. Importantly, all the patients had a pathological diagnosis of AD with Braak stage of V or VI (4). This means that all patients had NFT in the hippocampus by definition, and the method focused on separating the subset of patients with NFT predominantly in the hippocampus (limbic-predominant AD) versus the subset of patients with NFT predominantly in the cortical regions (hippocampal-sparing AD). Remainder of the patients had a rather balanced NFT count in the hippocampus and cortical regions, and were classified as typical AD.

Murray’s method (1) motivated many subsequent sMRI studies (see (2,40)). However, these studies rely on sMRI, a marker of unspecific neurodegeneration. This raises several problems. Firstly, while sMRI can reliably track neuropathologically-defined subtypes (47), the actual distribution of NFT in sMRI subtypes remains largely unknown. Recent studies have provided interesting preliminary data on tau PET uptake in sMRI-based subtypes (28,48). Secondly, the published sMRI subtype studies quite likely included patients in Braak stage IV for NFT or lower. Thirdly, all sMRI studies except for the study by Risacher et al. (6) investigated cohorts including both amyloid-beta positive and negative AD dementia patients, while all the patients in Murray et al. (1) had a pathological diagnosis of AD. Fourthly, neurodegeneration is downstream to NFT pathology (49), and there is a time gap until overt brain atrophy can be visually observed or captured by automatic methods for data analysis. Nonetheless, some data-driven methods may capture subtle differences in regional covariance in the absence of overt brain atrophy, mitigating this problem. Altogether, we still need to get a better understanding of the correspondence between neuropathologically-, sMRI-, and tau PET-defined subtypes. A major contribution of our current study is that subtypes identified with sMRI and tau PET are not interchangeable at the individual level.

At the group level, findings for the demographic and clinical measures were in agreement with previously reported studies and a recent meta-analysis (2). Broadly, typical AD was the most frequent subtype; typical and limbic-predominant AD were older in comparison to the hippocampal-sparing and minimal atrophy AD; MMSE scores were mostly comparable across subtypes with minimal atrophy AD showing the highest scores; a lower proportion of *APOE* ε4 carriers belonged to hippocampal-sparing relative to typical and limbic-predominant AD; and hippocampal-sparing AD had the highest levels of education.

The head-to-head results are best understood by considering individual exemplars. A consistent scenario is RID 2239: across the sMRI methods, this individual was classified as hippocampal-sparing or minimal atrophy AD whereas across the tau PET methods, the individual was classified as typical AD. The difference in sMRI-based subtyping could be attributed to differences in cut points for abnormality across methods. The fact that the corresponding tau PET-based subtype was typical AD (higher severity) could suggest greater tau pathology relative to structural atrophy. A more challenging case is RID 6377: across the sMRI methods, this individual was classified as typical, hippocampal-sparing, limbic-predominant, or minimal atrophy AD, whereas across the tau PET-based method, the individual was classified as limbic-predominant AD. Some differences in sMRI-based subtyping are relatively more plausible than others, considering the above-mentioned typicality and severity dimensions (2). To instantiate, it may be plausible that this individual demonstrated typical AD (with one method (6)) and limbic-predominant AD (with another method (7)), as these two subtypes are close to each other along the typicality dimension (2). However, classification as limbic-predominant AD (with one method (7)) and hippocampal-sparing AD (with another method (5)) seem incompatible, since these two subtypes correspond to the extremities of the typicality dimension. Therefore, a classification with all four subtypes for the same individual leaves the case biologically uninterpretable and claims for consensus in the field as we aim for precision medicine.

Despite having several caveats, these previous studies have made important contributions. Byun et al. (5) and Risacher et al. (6) translated the NFT-based method by Murray et al. (1) to sMRI data, and Charil et al. (25) translated the method to tau PET. Our analyses of methodological variations showed that the age correction made a stronger impact on agreements among methods, as compared with the number of cortical regions or the PVC. That impact was more prominent for sMRI-based methods than for tau PET-based methods; and for limbic-predominant and hippocampal-sparing subtypes than for typical AD. The contribution of aging to hippocampal atrophy may be at the basis of this finding. The lower disagreement in typical AD relative to the other subtypes is akin to the diagnostic challenge in the clinical setting. An interesting result of our study is that the method of adjustment (single model for all covariates *vs*. separate model for each covariate) increased the disagreement. Future studies should take this finding into account when deciding on how to correct for potential confounders.

Ongoing research is moving the field forward by characterization of subtypes not only at the stage of AD dementia but also at earlier stages such as prodromal AD (50,51). Preliminary data show that such characterization could be extended and evaluated at even the earliest stages of preclinical AD or individuals with subjective cognitive decline (52). It could be speculated that relative to full-blown dementia, atrophy levels are likely modest even if there exists overt tau pathology at pre-dementia stages. This could result in a greater dissociation between atrophy and tau pathology, in turn, leading to lower agreement across subtyping methods. In this scenario, understanding of profiles at the group-level alone is insufficient. Agreement at the individual level is thus warranted and lack thereof will prevent or delay the use of subtyping in clinical routine, clinical trials, and research. Therefore, there is an urgent need for harmonization of the different subtyping methods. To this end, we claim for establishing a framework for benchmarking for future studies. A possibility could be selection of a well-characterized cohort (preferably with multimodal data including postmortem and antemortem data in a longitudinal setting) and establishment of metrics used to evaluate the performance of the subtypes methods (e.g. group-level and individual-level results). The dataset should be standard so that it can be utilized by future subtyping methods to ensure individual-level consistency across methods. The dataset should also be open and accessible to all researchers in the field. As a preliminary step, we provide all the data used for subtyping in this study along with ADNI RIDs (see **Supplemental Data File**).

This study has some limitations. The cohort was part of the ADNI, which has strict selection criteria and excludes individuals with non-amnestic presentations or cerebrovascular pathology. It is likely that agreement among subtyping methods is different in clinically oriented or more heterogeneous cohorts. Hypothesis-driven methods are well covered in our study (1,5–7,25). However, previous subtyping studies have applied many different data-driven methods. We selected Poulakis’ method (15) and our current study cannot provide direct insight on methods used by other groups (2,40). However, the selection of subtyping methods illustrates the case made in the current study. We based our analyses on cross-sectional tau PET and sMRI data. The next step should be to include longitudinal data. However, the availability of such a dataset is limited at present, particularly for tau PET. Longitudinal data will be relevant to investigate disease progression in the subtypes, disentangling the disagreement due to the temporal lag between NFT accumulation (tau PET) and brain atrophy (sMRI) from pure methodological noise.

The field of biological subtypes is expanding rapidly with investigation of multiple modalities/biomarkers and extending to pre-dementia stages and other neurodegenerative diseases (40). We conclude that subtyping methods may appear comparable across studies, at the group-level. However, a major finding of the present study is the large disagreement among subtyping methods at the individual level. Hence, there is an urgent need for consensus and harmonization across subtyping methods. To achieve this, we suggest establishment of an accessible and standard framework for benchmarking. A comprehensive dataset along with clear evaluation metrics will facilitate a fair comparison and ultimately ensure better agreement among future subtyping methods.

## Data Availability

All data used in preparation of this manuscript is made available in the supplementary materials.

http://adni.loni.usc.edu/

## ACKNOWLEDGEMENTS

This study was funded by the Swedish Foundation for Strategic Research (SSF); the Strategic Research Programme in Neuroscience at Karolinska Institutet (StratNeuro); the Swedish Research Council (VR, 2016-02282); the regional agreement on medical training and clinical research (ALF) between Stockholm County Council and Karolinska Institutet; Center for Innovative Medicine (CIMED); the Swedish Alzheimer Foundation; the Swedish Brain Foundation; the Åke Wiberg Foundation; Demensfonden; Stiftelsen Olle Engkvist Byggmästare; and Birgitta och Sten Westerberg. The funding sources did not have any involvement on the study design; collection, analysis, and interpretation of data; writing of the report; and the decision to submit the article for publication.

Data collection and sharing for this study was funded by the Alzheimer’s Disease Neuroimaging Initiative (ADNI) (National Institutes of Health Grant U01 AG024904) and DOD ADNI (Department of Defense award number W81XWH-12-2-0012). ADNI is funded by the National Institute on Aging, the National Institute of Biomedical Imaging and Bioengineering, and through generous contributions from the following: Alzheimer’s Association; Alzheimer’s Drug Discovery Foundation; BioClinica, Inc.; Biogen Idec Inc.; Bristol-Myers Squibb Company; Eisai Inc.; Elan Pharmaceuticals, Inc.; Eli Lilly and Company; F. Hoffmann-La Roche Ltd and its affiliated company Genentech, Inc.; GE Healthcare; Innogenetics, N.V.; IXICO Ltd.; Janssen Alzheimer Immunotherapy Research & Development, LLC.; Johnson & Johnson Pharmaceutical Research & Development LLC.; Medpace, Inc.; Merck & Co., Inc.; Meso Scale Diagnostics, LLC.; NeuroRx Research; Novartis Pharmaceuticals Corporation; Pfizer Inc.; Piramal Imaging; Servier; Synarc Inc.; and Takeda Pharmaceutical Company. The Canadian Institutes of Health Research is providing funds to support ADNI clinical sites in Canada. Private sector contributions are facilitated by the Foundation for the National Institutes of Health (www.fnih.org). The grantee organization is the Northern California Institute for Research and Education, and the study is coordinated by the Alzheimer’s Disease Cooperative Study at the University of California, San Diego. ADNI data are disseminated by the Laboratory for Neuro Imaging at the University of California, Los Angeles.

Data used in this study were obtained from the Alzheimer’s Disease Neuroimaging Initiative (ADNI) database (adni.loni.ucla.edu). As such, the investigators within the ADNI contributed to the design and implementation of ADNI and/or provided data but did not participate in the analysis or writing of this report. A complete listing of ADNI investigators can be found at: http://adni.loni.ucla.edu/wp-content/uploads/how_to_apply/ADNI_Acknowledgement_List.pdf.

## AUTHOR DECLARATIONS

The relevant ethical guidelines have been followed. All necessary IRB and/or ethics committee approvals were obtained, and details of the IRB/oversight body have been included in the manuscript.

## REFERENCES

1. Murray ME, Graff-Radford NR, Ross OA, Petersen RC, Duara R, Dickson DW. Neuropathologically defined subtypes of Alzheimer’s disease with distinct clinical characteristics: a retrospective study. Lancet Neurol. 2011;10(9):785–96.

2. Ferreira D, Nordberg A, Westman E. Biological subtypes of Alzheimer disease: A systematic review and meta-analysis. Neurology. 2020;

3. Whitwell JL, Graff-Radford J, Tosakulwong N, Weigand SD, Machulda M, Senjem ML, et al. [18F] AV-1451 clustering of entorhinal and cortical uptake in Alzheimer’s disease. Ann Neurol. 2018;83(2):248–57.

4. Braak H, Braak EVA. Staging of Alzheimer’s disease-related neurofibrillary changes. Neurobiol Aging. 1995;16(3):271–8.

5. Byun MS, Kim SE, Park J, Yi D, Choe YM, Sohn BK, et al. Heterogeneity of regional brain atrophy patterns associated with distinct progression rates in Alzheimer’s disease. PLoS One. 2015;10(11):e0142756.

6. Risacher SL, Anderson WH, Charil A, Castelluccio PF, Shcherbinin S, Saykin AJ, et al. Alzheimer disease brain atrophy subtypes are associated with cognition and rate of decline. Neurology. 2017;89(21):2176–86.

7. Ferreira D, Verhagen C, Hernández-Cabrera JA, Cavallin L, Guo C-J, Ekman U, et al. Distinct subtypes of Alzheimer’s disease based on patterns of brain atrophy: longitudinal trajectories and clinical applications. Sci Rep. 2017;7:46263.

8. Ferreira D, Shams S, Cavallin L, Viitanen M, Martola J, Granberg T, et al. The contribution of small vessel disease to subtypes of Alzheimer’s disease: a study on cerebrospinal fluid and imaging biomarkers. Neurobiol Aging. 2018;70:18–29.

9. Ferreira D, Pereira JB, Volpe G, Westman E. Subtypes of Alzheimer’s Disease Display Distinct Network Abnormalities Extending Beyond Their Pattern of Brain Atrophy. Front Neurol. 2019;10:524.

10. Persson K, Eldholm RS, Barca ML, Cavallin L, Ferreira D, Knapskog A-B, et al. MRI-assessed atrophy subtypes in Alzheimer’s disease and the cognitive reserve hypothesis. PLoS One. 2017;12(10).

11. Oppedal K, Ferreira D, Cavallin L, Lemstra AW, Ten Kate M, Padovani A, et al. A signature pattern of cortical atrophy in dementia with Lewy bodies: a study on 333 patients from the European DLB consortium. Alzheimer’s Dement. 2019;15(3):400–9.

12. Machado A, Ferreira D, Grothe MJ, Eyjolfsdottir H, Almqvist PM, Cavallin L, et al. The cholinergic system and treatment response in subtypes of Alzheimer’s disease. medRxiv. 2020;

13. Ekman U, Ferreira D, Westman E. The A/T/N biomarker scheme and patterns of brain atrophy assessed in mild cognitive impairment. Sci Rep. 2018;8(1):1–10.

14. Ferreira D, Cavallin L, Larsson E, Muehlboeck J, Mecocci P, Vellas B, et al. Practical cut-offs for visual rating scales of medial temporal, frontal and posterior atrophy in A lzheimer’s disease and mild cognitive impairment. J Intern Med. 2015;278(3):277–90.

15. Poulakis K, Pereira JB, Mecocci P, Vellas B, Tsolaki M, Kłoszewska I, et al. Heterogeneous patterns of brain atrophy in Alzheimer’s disease. Neurobiol Aging. 2018;65:98–108.

16. Poulakis K, Ferreira D, Pereira JB, Smedby O, Vemuri P, Westman E. Fully Bayesian longitudinal unsupervised learning for the assessment and visualization of AD heterogeneity and progression. bioRxiv. 2019;854356.

17. Noh Y, Jeon S, Lee JM, Seo SW, Kim GH, Cho H, et al. Anatomical heterogeneity of Alzheimer disease: based on cortical thickness on MRIs. Neurology. 2014;83(21):1936–44.

18. Na HK, Kang DR, Kim S, Seo SW, Heilman KM, Noh Y, et al. Malignant progression in parietal-dominant atrophy subtype of Alzheimer’s disease occurs independent of onset age. Neurobiol Aging. 2016;47:149–56.

19. Hwang J, Kim CM, Jeon S, Lee JM, Hong YJ, Roh JH, et al. Prediction of Alzheimer’s disease pathophysiology based on cortical thickness patterns. Alzheimer’s Dement Diagnosis, Assess Dis Monit. 2016;2:58–67.

20. Dong A, Honnorat N, Gaonkar B, Davatzikos C. CHIMERA: Clustering of heterogeneous disease effects via distribution matching of imaging patterns. IEEE Trans Med Imaging. 2015;35(2):612–21.

21. Dong A, Toledo JB, Honnorat N, Doshi J, Varol E, Sotiras A, et al. Heterogeneity of neuroanatomical patterns in prodromal Alzheimer’s disease: links to cognition, progression and biomarkers. Brain. 2017;140(3):735–47.

22. Varol E, Sotiras A, Davatzikos C, Initiative ADN. HYDRA: Revealing heterogeneity of imaging and genetic patterns through a multiple max-margin discriminative analysis framework. Neuroimage. 2017;145:346–64.

23. Zhang X, Mormino EC, Sun N, Sperling RA, Sabuncu MR, Yeo BTT, et al. Bayesian model reveals latent atrophy factors with dissociable cognitive trajectories in Alzheimer’s disease. Proc Natl Acad Sci. 2016;113(42):E6535–44.

24. Park J-Y, Na HK, Kim S, Kim H, Kim HJ, Seo SW, et al. Robust identification of Alzheimer’s disease subtypes based on cortical atrophy patterns. Sci Rep. 2017;7(1):1–14.

25. Charil A, Shcherbinin S, Southekal S, Devous MD, Mintun M, Murray ME, et al. Tau Subtypes of Alzheimer’s Disease Determined in vivo Using Flortaucipir PET Imaging. J Alzheimer’s Dis. 2019;(Preprint):1–12.

26. Marinescu R V, Eshaghi A, Lorenzi M, Young AL, Oxtoby NP, Garbarino S, et al. DIVE: A spatiotemporal progression model of brain pathology in neurodegenerative disorders. Neuroimage. 2019;192:166–77.

27. Young AL, Marinescu R V, Oxtoby NP, Bocchetta M, Yong K, Firth NC, et al. Uncovering the heterogeneity and temporal complexity of neurodegenerative diseases with Subtype and Stage Inference. Nat Commun. 2018;9(1):1–16.

28. Jeon S, Kang JM, Seo S, Jeong HJ, Funck TF, Lee S-Y, et al. Topographical Heterogeneity of Alzheimer’s Disease Based on MR Imaging, Tau PET, and Amyloid PET. Front Aging Neurosci. 2019;11:211.

29. Mueller SG, Weiner MW, Thal LJ, Petersen RC, Jack C, Jagust W, et al. The Alzheimer’s disease neuroimaging initiative. Neuroimaging Clin. 2005;15(4):869–77.

30. Shaw LM, Vanderstichele H, Knapik-Czajka M, Clark CM, Aisen PS, Petersen RC, et al. Cerebrospinal fluid biomarker signature in Alzheimer’s disease neuroimaging initiative subjects. Ann Neurol. 2009;65(4):403–13.

31. Joshi AD, Pontecorvo MJ, Clark CM, Carpenter AP, Jennings DL, Sadowsky CH, et al. Performance characteristics of amyloid PET with florbetapir F 18 in patients with Alzheimer’s disease and cognitively normal subjects. J Nucl Med. 2012;53(3):378–84.

32. Muehlboeck J, Westman E, Simmons A. TheHiveDB image data management and analysis framework. Front Neuroinform. 2014;7:49.

33. Desikan RS, Ségonne F, Fischl B, Quinn BT, Dickerson BC, Blacker D, et al. An automated labeling system for subdividing the human cerebral cortex on MRI scans into gyral based regions of interest. Neuroimage. 2006;31(3):968–80.

34. Destrieux C, Fischl B, Dale A, Halgren E. Automatic parcellation of human cortical gyri and sulci using standard anatomical nomenclature. Neuroimage. 2010;53(1):1–15.

35. Fischl B, Salat DH, Busa E, Albert M, Dieterich M, Haselgrove C, et al. Whole brain segmentation: automated labeling of neuroanatomical structures in the human brain. Neuron. 2002;33(3):341–55.

36. Mårtensson G, Ferreira D, Cavallin L, Muehlboeck J-S, Wahlund L-O, Wang C, et al. AVRA: Automatic visual ratings of atrophy from MRI images using recurrent convolutional neural networks. NeuroImage Clin. 2019;23:101872.

37. Mårtensson G, Ferreira D, Granberg T, Cavallin L, Oppedal K, Padovani A, et al. The reliability of a deep learning model in clinical out-of-distribution MRI data: a multicohort study. arXiv Prepr arXiv191100515. 2019;

38. Greve DN, Salat DH, Bowen SL, Izquierdo-Garcia D, Schultz AP, Catana C, et al. Different partial volume correction methods lead to different conclusions: an 18F-FDG-PET study of aging. Neuroimage. 2016;132:334–43.

39. Rousset, OG. Correction for partial volume effects in PET?: principle and validation. J Nucl Med [Internet]. 1998 [cited 2020 Feb 18];39:904–11. Available from: http://ci.nii.ac.jp/naid/10025136344/en/

40. Habes M, Grothe MJ, Tunc B, McMillan C, Wolk DA, Davatzikos C. Disentangling heterogeneity in Alzheimer’s disease and related dementias using data-driven methods. Biol Psychiatry. 2020;

41. Lemoine L, Leuzy A, Chiotis K, Rodriguez-Vieitez E, Nordberg A. Tau positron emission tomography imaging in tauopathies: the added hurdle of off-target binding. Alzheimer’s Dement Diagnosis, Assess Dis Monit. 2018;10:232–6.

42. Lee CM, Jacobs HIL, Marquié M, Becker JA, Andrea N V, Jin DS, et al. 18F-Flortaucipir binding in choroid plexus: related to race and Hippocampus signal. J Alzheimer’s Dis. 2018;62(4):1691–702.

43. Schöll M, Lockhart SN, Schonhaut DR, O’Neil JP, Janabi M, Ossenkoppele R, et al. PET imaging of tau deposition in the aging human brain. Neuron. 2016;89(5):971–82.

44. Lowe VJ, Curran G, Fang P, Liesinger AM, Josephs KA, Parisi JE, et al. An autoradiographic evaluation of AV-1451 Tau PET in dementia. Acta Neuropathol Commun. 2016;4(1):58.

45. Ikonomovic MD, Abrahamson EE, Price JC, Mathis CA, Klunk WE. [F-18] AV-1451 positron emission tomography retention in choroid plexus: More than “off-target” binding. Ann Neurol. 2016;80(2):307–8.

46. Landis JR, Koch GG. The measurement of observer agreement for categorical data. Biometrics. 1977;159–74.

47. Whitwell JL, Dickson DW, Murray ME, Weigand SD, Tosakulwong N, Senjem ML, et al. Neuroimaging correlates of pathologically defined subtypes of Alzheimer’s disease: a casecontrol study. Lancet Neurol. 2012;11(10):868–77.

48. Ossenkoppele R, Lyoo CH, Sudre CH, van Westen D, Cho H, Ryu YH, et al. Distinct tau PET patterns in atrophy-defined subtypes of Alzheimer’s disease. Alzheimer’s Dement. 2019;

49. Dubois B, Feldman HH, Jacova C, Hampel H, Molinuevo JL, Blennow K, et al. Advancing research diagnostic criteria for Alzheimer’s disease: the IWG-2 criteria. Lancet Neurol. 2014;13(6):614–29.

50. Machulda MM, Lundt ES, Albertson SM, Kremers WK, Mielke MM, Knopman DS, et al. Neuropsychological subtypes of incident mild cognitive impairment in the Mayo Clinic Study of Aging. Alzheimer’s Dement. 2019;15(7):878–87.

51. ten Kate M, Dicks E, Visser PJ, van der Flier WM, Teunissen CE, Barkhof F, et al. Atrophy subtypes in prodromal Alzheimer’s disease are associated with cognitive decline. Brain. 2018;141(12):3443–56.

52. Jung N-Y, Seo SW, Yoo H, Yang J-J, Park S, Kim YJ, et al. Classifying anatomical subtypes of subjective memory impairment. Neurobiol Aging. 2016;48:53–60.

